# Heterogeneous human exposure to arbovirus vectors in an African urban context using immuno-epidemiological biomarker of *Aedes aegypti* bites

**DOI:** 10.1101/2025.03.25.25324614

**Authors:** Bi Zamblé Hubert Zamble, Akré Maurice Adja, André Barembaye Sagna, François Dipomin Traore, Mintokapieu Didier Stéphane Kpan, Négnorogo Guindo-Coulibaly, Affoué Mireille Nadia Kouadio, Konan Rodolphe Mardoché Azongnibo, Danielle Dounin Zoh, Anne Poinsignon, Florence Fournet, Françoise Mathieu-Daude, Franck Remoue

## Abstract

Uncontrolled urbanization led to a specific environment, promoting the spread of *Aedes* mosquitoes in many African cities. The objective of the study was to assess human exposure to *Aedes* mosquito bites in the urban area of Abidjan, Côte d’Ivoire, by using antibody-based biomarkers. A cross-sectional study was undertaken during the short rainy season in 3 different neighborhoods (Bromakote, Anoumabo, and Petit-Bassam) presenting various socio-environmental and entomological characteristics. The specific IgG responses to *Aedes* Nterm-34kDa salivary peptide, previously validated as a relevant individual biomarker of exposure to mosquito bites, were assessed in children and analyzed according to neighborhoods and age classes. The specific IgG level was significantly different according to the 3 neighborhoods and higher in Bromakote compared to Anoumabo and Petit-Bassam. No significant difference in specific IgG level was observed according to age. We also noticed an association between the level of specific IgG responses and *Ae. aegypti* densities or various socio-environmental factors. This study indicated that Human exposure to *Aedes* vector appeared to be dependent on neighborhoods within the same city, which could be related to some socio-environmental factors. Antibody-based biomarkers of human exposure to *Aedes* bites could be a helpful tool for assessing the heterogeneity of urban exposure to arbovirus vectors in the framework of monitoring the risk of arbovirus infections, and for evaluating the effectiveness of vector control strategies in national control programs.

## Introduction

Emerging and re-emerging diseases such as arboviral infections (dengue, yellow fever, chikungunya, and Zika) remain major health problems worldwide (Weetman *et al*., 2018). It is estimated that 2.5 billion people are at risk of dengue infection in the world. Around 200 000 people are affected each year by yellow fever infection around the world (Weetman *et al*., 2018). *Aedes aegypti* and *Aedes albopictus* mosquitoes, the major vectors of arboviruses, are worldwide spread, especially in urban settings (Vega-Rua *et al*., 2014).

In Africa, 43% of the population live in urban settings leading to the rapid and uncontrolled urbanization of many African cities (UN, 2018). Uncontrolled urbanization is often accompanied by many issues on the spread of vector-borne diseases (Uttara *et al*., 2012). This urbanization, driven by the expansion of African cities, creates favorable breeding sites for mosquito proliferation and potential emergence (Li *et al*., 2014). Among them, *Aedes aegypti* is the major *Aedes* species in urban areas in African cities (Li *et al*., 2014; Fofana *et al*., 2019).

During the last decades, many epidemics of arboviral diseases occurred in the main cities of some African countries, as observed in West Africa: in Senegal in 2009 and on major in Ouagadougou, Burkina Faso in 2023 (Faye *et al*., 2014; WHO, 2016; Manigart *et al*., 2024). In Côte d’Ivoire, Abidjan is the largest and most densely populated city and a prime example of an expanding African megalopolis with uncontrolled urbanization. In this city, 192 cases of dengue fever were recorded during the 2017 outbreak. Although poorly documented, yellow fever transmission occurred in Côte d’Ivoire and some confirmed (n=5) cases were also recorded in the northern part of the country in 2011 (WHO, 2017 and WHO, 2011).

No vaccine or medicines are available to control arboviral diseases, except for two efficient vaccines against yellow fever and Japanese encephalitis. The most effective measures against these diseases are therefore prevention and vector population control. That involves conventional methods such as personal protection, community mobilization, insecticide treatments (Andersson *et al*., 2015), and innovative approaches such as sterile insect techniques, release of *Wolbachia*-infected *Aedes* (Tur *et al*., 2021) or biological control of *Aedes* populations (Huang *et al*., 2017; Utarini *et al*. 2021).

The evaluation of arbovirus transmission risk and vector control effectiveness is crucial. Regarding *Aedes* vectors, *Stegomyia* indices such as House Index (HI), Container Index (CI), and Breteau Index (BI) are calculated and used as indicators for the risk of arbovirus transmission (Bowman *et al*., 2014). These parameters present several limitations because they represent only an indirect proxy of the *Aedes* adult density in a study site, and do not directly measure the human-vector contact (Boyer *et al*., 2014). In addition, these entomological methods assess human exposure to vector bites at the population level and do not consider the heterogeneity of inter-individual exposure. Therefore, WHO underlines the need for the development of alternative indicators to accurately evaluate the real and direct human exposure to *Ae. aegypti* mosquito bites and to estimate the potential risk of *Aedes aegypti* borne pathogen transmission in an exposed population (WHO, 2014). During blood-feeding, concomitantly to blood intake, female mosquitoes inject saliva containing a cocktail of biologically active proteins that counteract host homeostasis and modulate the vertebrate immune response (Ribeiro *et al*., 2003). The injection of vector saliva can also elicit a human antibody (Ab) response against the salivary proteins (Doucoure *et al*., 2015; Drame *et al*., 2010). An interesting approach exploits the immunological properties of mosquito saliva to identify immuno-epidemiological biomarkers of human exposure to mosquito bites (Fontaine *et al*., 2011, Doucoure *et al*., 2012). To avoid immune cross-reactivity between arthropods and the limitations of whole saliva use, the identification of antigenic salivary proteins or peptides specific to vector species/genus was achieved and demonstrated. Specific immunoglobulin G (IgG) antibody responses have been validated as a serological biomarker of exposure to the main mosquito vectors such as *An. gambiae* (IgG response to gSG6 or CE5 salivary proteins, or to the gSG6-P1 salivary peptide), the vector of human malaria *Plasmodium* (Poinsignon *et al*., 2008; Rizzo *et al*., 2014). In the case of arboviral diseases, several studies have validated the IgG Ab response to only one *Aedes* salivary peptide (the *Ae. aegypti* Nterm-34 kDa peptide) as a biomarker of human exposure to *Aedes* genus bites, and thus an indicator for evaluating the risk of arbovirus transmission, and as a relevant tool to assess vector control strategies (Elanga Ndille *et al*., 2012; Sagna *et al*., 2019; Sagna *et al*., 2018). In a study in Vientiane city, Laos, the human IgG response against this salivary peptide showed an inter-individual heterogeneity of exposure to *Ae. aegypti* bites in semi-urban and urban settings (Elanga Ndille *et al*., 2014). These results suggested also that the IgG responses can identify areas at risk of dengue transmission (Elanga Ndille *et al*., 2014). In another study, the human IgG response to this salivary peptide has been validated as a relative quantitative indicator to evaluate the efficacy of vector control strategies (Elanga Ndille *et al*., 2016). This study highlighted that IgG response to salivary peptide presented a short-life property with no cumulative Ab response, and a rapid decreased (2 to 4 weeks) of specific IgG level was observed after the stop of human-*Aedes* contact. This present study aimed to use the IgG responses to *Ae. aegypti* Nterm-34 kDa salivary peptide to evaluate the urban heterogeneity of human exposure to the major arbovirus vectors, *Ae. aegypti* in Abidjan city, Côte d’Ivoire.

## Material and methods

### Study sites

The study was conducted in three urban settings of Abidjan city (Fig.1), the largest and the most populated city (about 5 million inhabitants) in Côte d’Ivoire. Located in the southern region and crossed by Ebrie lagoon, Abidjan city is characterized by a sub-equatorial climate, hot and humid, which includes a main rainy season (May to July), a short rainy season (September to November), separated by two dry seasons. Rainfall in Abidjan is abundant with more than 1,500 mm per year. The temperature is around 27°C all year round and the average annual humidity is over 80 %. During the short rainy season, covered by the present study, the average temperature was 26°C and the average rainfall 123 mm over 3 months (Kpan *et al*., 2021).

**Figure 1.**
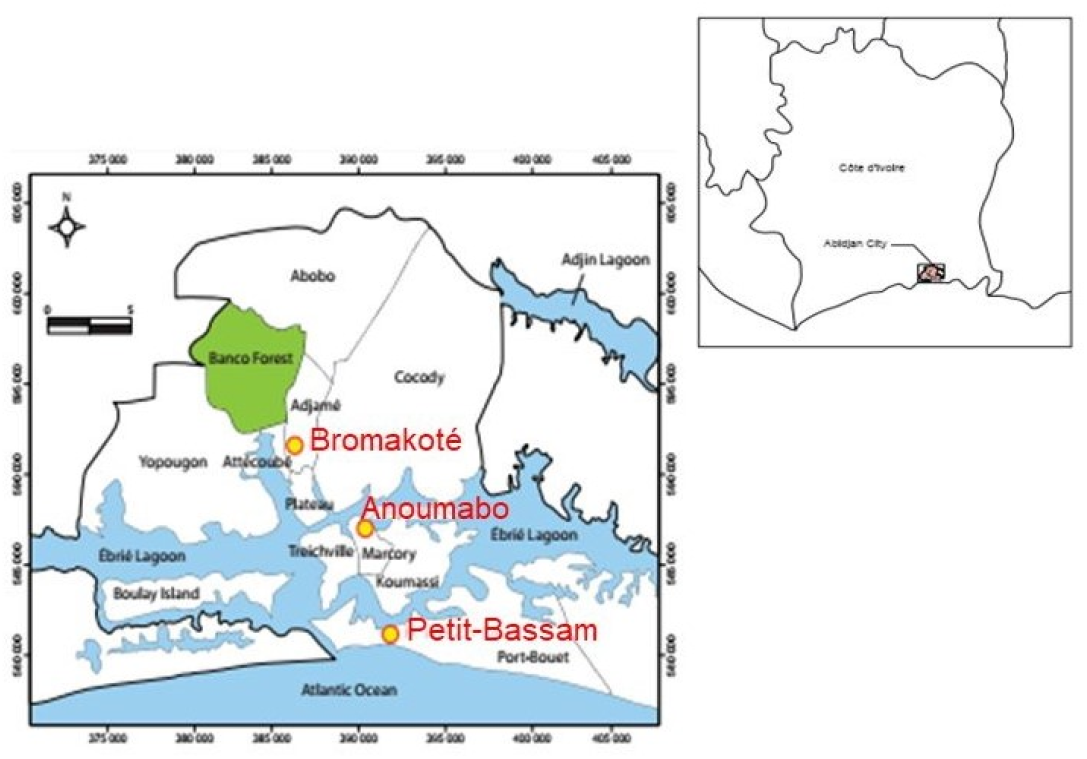
Map of the study area in the city of Abidjan, Côte d’Ivoire. The different study sites (Bromakote, Anoumabo, Petit-Bassam) are indicated by yellow dots.

Three municipalities (Adjame, Marcory and Port-Bouët) of Abidjan were selected and one neighborhood in each municipality was included in the study. In Adjame, the Bromakote neighborhood was selected because of its extensive commercial activities and the numerous exchanges of people (transport) between the different neighborhoods across Abidjan and between African countries due to the location of the main bus station of the city. This neighborhood also includes the railway station that connects Côte d’Ivoire to Burkina Faso for people and goods. The choice of Anoumabo in Marcory municipality was based on its precarious nature and therefore its environment potentially favorable to the development of mosquitoes. The choice of Petit-Bassam, in Port-Bouet municipality, was driven by the presence of the autonomous port in this neighborhood, resulting in significant traffic of goods and the potential introduction of vector species and pathogens. Besides, the presence of *Ae. albopictus*, a major vector of arbovirus, and cases of dengue infections have already been reported in this neighborhood (Konan *et al*., 2013).

In the objective to describe the socio-environmental characteristics of the neighborhoods in relation to the risk of proliferation of *Aedes* vector, one hundred households were randomly selected in each neighborhood. The Geographic Information System software (version QGIS 2.14.4) was used to randomly select households. Information on house type, building materials and social practices (water storage, domestic waste and wastewater management, type of toilets) were collected in the three neighborhoods.

### Study population

The entire study was carried out on a population of 880 children aged from 6 months to 14 years, enrolled at the school site in each neighborhood, in parallel to a malaria survey on this target age population. This cross-sectional study was performed during the short rainy season (SRS) from September to November 2015. The present immunological study was conducted on a sub-sample of the initial cohort and included 436 children (Table 1). For the immunological assays, blood samples were collected in November 2015 during the SRS, at the fingertips in microtainer tubes (Sarsted, Marnay, France) and serum was obtained, fractionated into aliquots and then frozen at -20 ° C until used.

**Table 1.** Socio-environmental characteristics of the study sites

|  | Neighborhoods |  |  |  |
| --- | --- | --- | --- | --- |
|  | Anoumabo | Bromakote | Petit Bassam | p-value |
| Neighborhood characteristics n (%)* |  |  |  |  |
| House type | n.a= 55 | n.a= 87 | n.a= 98 |  |
| Common house | 48 (87.28) | 74 (83.15) | 89 (90.82) | NS |
| Apartment | 03 (05.45) | 11 (12.36) | 02 (02.04) | 0.01 |
| Individual house | 04 (07.27) | 02 (02.25) | 06 (06.12) | NS |
| Construction materials | n.a= 45 | n.a=89 | n.a= 80 |  |
| Cement | 38 (84.44) | 83 (93.26) | 72 (73.47) | NS |
| Wood | 04 (08.89) | 05(05.62) | 08 (08.16) | NS |
| Improved banco | 02 (04.44) | 01 (01.12) | 0 | NS |
| Banco (simple and improved) | 01 (02.22) | 0 | 0 | NS |
| Water storage containers | n.a = 48 | n.a=74 | n.a= 75 |  |
| Uncovered containers | 40 (83.33) | 58 (78.38) | 57 (76.00) | 0.001 |
| Covered containers | 01 (02.08) | 01(01.35) | 10 (13.33) | 0.02 |
| Both covered and uncovered containers | 07 (14.59) | 15 (20.27) | 08 (10.67) | NS |
| Waste management | n.a = 50 | n.a= 87 | n.a=97 |  |
| Collected by companies | 17 (34.00) | 48 (55.17) | 24 (24.74) | 0.005 |
| Dropped off near the house | 09 (18.00) | 04 (04.60) | 60 (61.86) | 0.001 |
| Burned near the house | 01 (20.00) | 0 | 01 (01.03) | NS |
| Collected or dropped off near the house | 23 (73.00) | 35 (40.23) | 12 (12.37) | 0.001 |

|  | Neighborhoods |  |  | *p*-value |
| --- | --- | --- | --- | --- |
|  | Anoumabo | Bromakote | Petit Bassam |  |
| **Wastewater management** | **n.a = 55** | **n.a = 87** | **n.a= 98** |  |
| Septic pits | 21 (38.18) | 55 (63.22) | 16 (16.33) | <0.001 |
| Sewage channels | 05 (09.00) | 17 (19.54) | 04 (04.08) | 0.0001 |
| Septic pits or abandoned wells | 09 (16.36) | 07 (08.05) | 02 (02.04) | 0.006 |
| Pour in the street | 19 (14.55) | 05 (05.75) | 71 (72.45) | <0.001 |
| Septic pits or pour in the street | 01 (01.82) | 02 (02.30) | 05 (05.10) | NS |
| Sewage channels or abandoned wells | 0 | 01(01.15) | 0 | NS |
|  |  |  | 0 |  |
| **Toilet type** | **n.a = 55** | **n.a = 89** | **n.a= 94** |  |
| Modern toilet | 11 (20.00) | 15 (16.85) | 28 (28.57) | 0.097 |
| Latrine | 43 (78.18) | 72 (80.9) | 66 (67.35) | NS |
| Modern toilet and latrine | 01 (01.82) | 02 (02.25) | 0 | NS |

n.a: number of answers
\* All percentage calculated for each characteristic in corresponding column
NS : Not Significant between the 3 neighborhoods

The Kruskal-Wallis test on children’s age indicated a significant difference in the mean of children’s age between the studied neighborhoods (*p*=0.0002). Comparison two-by-two (Dunn’s test and adjusted with Bonferroni method) indicated firstly a significant difference in the age mean between children from Bromakote and those living in Petit Bassam, and then between those living in Anoumabo and Bromakote (p-adjusted=0.001 and p-adjusted= 0.0007 respectively).

### Aedes aegypti Nterm-34 kDa salivary peptide

The Nterm-34 kDa salivary peptide from *Ae. aegypti* was synthesized and purified (> 95 %) by Genepep SA (Saint Jean de Vedas, France). The peptide was shipped in lyophilized form and then suspended in milliQ water and frozen at -20°C until use.

### Evaluation of human IgG level to Nterm-34 kDa salivary peptide

IgG level to Nterm-34 kDa peptide was measured by Enzyme-Linked Immuno-Sorbent Assay (ELISA) as previously described (Elanga Ndille *et al*., 2012). Briefly, MaxiSorp flat-bottom 96-well (Nunc, Roskilde, Denmark) were coated with Nterm-34 kDa peptide at a final concentration of 20 µg/ml in coating buffer (PBS) and incubated for 150 min at 37°C. Plates were blocked with 200 μL of protein-free Blocking Buffer (Thermo Scientific, Rockford, USA) and incubated for 60 min at 37°C. Individual sera were diluted in buffer (PBS-Tween 1%) and incubated at a final dilution of 1/80. Monoclonal mouse biotinylated anti-human IgG (BD Pharmingen, San Diego, CA) was added in at a 1/500 dilution (PBS-Tween 1%) and incubated for 90 min at 37°C. Peroxidase-conjugated Streptavidin (GE Healthcare, Orsay, France) was then added (1/1000 in PBS-Tween 1%) and incubated for 60 min at 37°C. Colorimetric development was carried out with 2,2’-azino-bis ethylbenzothiazoline 6-sulfonic acid diammonium salt (ABTS, Sigma, Saint-Louis, MO, USA) in 50 mM citrate buffer (pH = 4 containing 0.003% H2O2) and absorbance (optical density [OD]) was measured at 405 nm (Multisakan GO Thermo Scientific). Each sample was tested in duplicate wells containing Nterm-34 kDa and in a well without antigen (Nterm-30 kDa) to measure non-specific reactions. Individual results were expressed as the ΔOD value: ΔOD = ODx − ODn, where ODx represents the mean of the individual OD value in both wells with salivary antigen and ODn the individual OD value in a blank well containing no antigen.

### Entomological data

In the present study, three entomological methods were used according to the mosquito stage as previously described (Kpan *et al*., 2021). The identification of breeding sites was conducted during larval survey while ovitraps (eggs were reared until the adult stage at insectarium) and sticky traps were used for adult mosquitoes evaluation.

Using the random sampling tool of the QGIS 2.14.4, 20 geographical points were generated within each of the three study sites. One ovitrap and one sticky trap were placed at each geographical point during the study period (1 month). As a result, 20 ovitraps and 20 sticky traps were used to collect *Aedes* eggs and *Ae. aegypti* adults during the studied season in each 3 studied sites. Data in this present study were collected during the SRS in November 2015.

*Ae. aegypti* larvae population was estimated through visiting 300 households (100 in each neighborhood) and by identifying potential breeding containers in each household. The collected larvae were reared in an insectarium and morphologically identified after emergence. The *Stegomyia* indices have been calculated for the risk of arboviruses transmission by *Ae. aegypti* according to WHO criteria: i) the House Index (HI) represents the percentage of houses infested by *Ae. aegypti* larvae; ii) the Container Index (CI) is the percentage of containers infested by *Ae. aegypti* larvae, and iii) the Bretau Index (BI) is the number of positive breeding sites in 100 visited houses. According to these *Stegomyia indices*, the WHO criteria indicate that: i) an area is considered to be at high risk of transmission of dengue and chikungunya viruses by *Aedes aegypti* if HI or BI or CI exceed 5, 20, and 3%, respectively, and ii) an area is considered to be at high risk of favoring the transmission of Zika and yellow fever viruses by *Aedes aegypti* if HI or BI or CI exceed 5, 4 and 3%, respectively (Kpan *et al*., 2021).

### Data analysis

Statistical analysis was done using Graph Pad Prism 5® software (San Diego, CA) and R (Version 3.5.3, R Core Team, Vienna, Austria). After checking that the data did not follow a Gaussian distribution (normal distribution), the non-parametric Mann-Whitney test was used to compare the Ab levels between two independent groups and the non-parametric Kruskal-Wallis test was used to compare the Ab levels between more than two groups. The Dunn’s post-hoc test was also performed for two by two comparisons in more than three groups, then the difference was adjusted with the Bonferroni method. The prop.test was used to compare proportions between two or more two groups. The chi-square test was used to compare the HI and CI between study sites. All differences were considered significant for *p*<0.05.

## Results

### Socio-environmental characteristics of neighborhoods

Environmental factors and sociological behaviors/practices (housing type, construction materials, water storage, domestic waste and waste water management) that could potentially be associated with human exposure to *Aedes* mosquitoes were analyzed and compared according to the three neighborhoods. A total of 55, 87, and 98 houses in Anoumabo, Bromakote, and Petit-Bassam, respectively, were randomly selected (among the 100 households by neighborhood of the entomological surve and visited allowing to identify common and divergent factors between the 3 neighborhoods (Table 1).

There was no clear difference in the housing type between the three neighborhoods, with the exception of the number of apartments, which was higher in Bromakote than in the two other sites (12.36 %; *p*=0.01). Most houses were constructed with cement and no significant difference in the construction materials was observed between the three sites.

Most residents in the three neighborhoods used the stored water for cooking or drinking, and the comparison of this practice (use of containers for water storage) was not statistically significant between neighborhoods (*p*=0.262, data not shown). The large majority of water storage containers found in all neighborhoods were uncovered (>75 %). However, a significant difference was observed in the proportion of uncovered water storage containers between neighborhoods (*p*=0.001) with higher proportion recorded in Anoumabo (83.33 %).

Regarding domestic waste management, a higher proportion of inhabitants in Bromakote declared that their waste was collected by private companies (55.17 %; *p*=0.005). In contrast, domestic waste was mostly dropped off near the house in Petit-Bassam (61.22 %; *p*<0.001). Fifty-five (55) inhabitants in Anoumabo, 80 in Bromakote, and 98 in Petit-Bassam provided information about wastewater management. Among them, a higher proportion in Bromakote (61.8 %) discharged wastewater into a septic pit compared to other neighborhoods (*p*<0.001). Channels were also used for discharging wastewater by 19.54 %, (*p*=0.006). Some inhabitants used both septic pit or abandoned wells with a higher proportion in Anoumabo (16.36 %; *p*<0.001). In addition, most people used latrine as a type of toilet, and a very low number of modern toilets was recorded in the three neighborhoods.

Altogether, the results indicated that some sociological practices such as wastewater and waste management, water storage mode and housing type appeared differently represented between the three neighborhoods.

### Specific IgG antibody level

IgG levels to Nterm-34 kDa salivary peptide were compared between the 3 studied neighborhoods in the sub-sample of 436 of children (Table 2, Fig. 2).

**Table 2.** Population characteristics and median of IgG level to Nterm-34 kDa salivary peptide in the study sites

|  | Neighborhoods |  |  | Total | p-value |
| --- | --- | --- | --- | --- | --- |
|  | Anoumabo | Bromakote | Petit Bassam |  |  |
| **Demographic data** |  |  |  |  |  |
| n (%) | 135 (30.96) | 122 (27.98) | 179 (41.06) | 436 |  |
| Age mean (95% CI) | 6.50 (5.95-7.06) | 4.98 (4.33-5.64) | 5.90 (6.43-6.96) | 6.05 (5.72-6.40) | 0.001 |
| **Age groups [years] (%)** |  |  |  |  |  |
| [1-5] | 54 | 73 | 78 |  | NS |
| [6-10] | 66 | 34 | 70 |  |  |
| [11-14] | 15 | 15 | 30 |  |  |
| **Gender ratio (Male/Female)** | 0.82 (61/74) | 0.92 (56/61) | 0.69 (73/105) | 0.80 (190/240) | 0.016 |
| **Anti-Nterm-34 kDa IgG median level (95% CI)** | 0.174 [0.118-0.271] | 0.321 [0.261-0.378] | 0.138 [0.106-0.173] |  | 0.001 |
| **Anti-Nterm-34 kDa IgG median level by age group (95% CI)** |  |  |  |  |  |
| [1-5] | 0.15 [0.16-0.34] | 0.35 [0.33-0.46] | 0.17 [0.19-0.28] | 0.21[0.27-0.33] | 0.001 |
| [6-10] | 0.20 [0.22-0.34] | 0.26 [0.25-0.54] | 0.12 [0.14-0.26] | 0.16[0.22-0.31] | 0.001 |
| [11-14] | 0.17 [0.14-0.35] | 0.20 [0.14-0.37] | 0.15 [0.12-0.26] | 0.16 [0.17-0.27] | NS |

NS : Not Significant between the 3 neighborhoods

**Figure 2.**
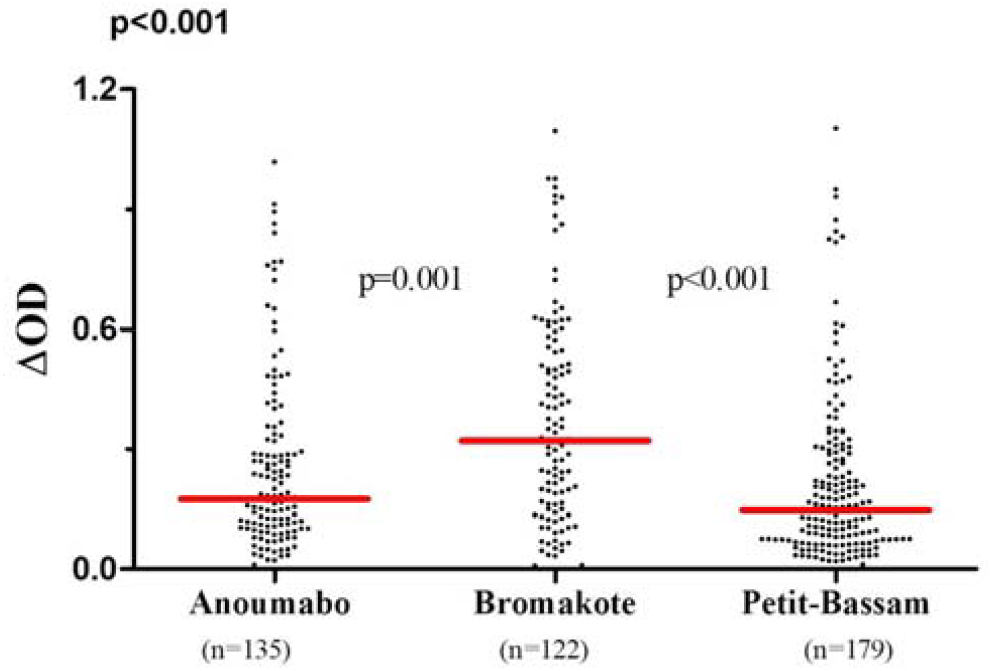
IgG responses to *Aedes aegypti* Nterm-34 kDa salivary peptide in the study sites. Each dot represents an individual IgG response (ΔOD) against the *Ae. aegypti* Nterm-34 kDa salivary peptide, and the horizontal bar indicates the median value of specific IgG levels for the site under study. The number of processed sera is indicated below (n).

First, high inter-individual heterogeneity of specific IgG responses (ΔOD ranging from 0 to 1.2) was observed within each neighborhood. Secondly, the level of specific IgG responses varied significantly between neighborhoods (*p<*0.001). These IgG levels were significantly higher in children from Bromakote than those from Anoumabo and Petit-Bassam (*p*<0.001). No significant difference in specific IgG levels was observed between Anoumabo and Petit-Bassam (*p*=0.102).

The median of IgG level was also compared between the 3 pre-defined age groups ([1-5], [6-10], and [11-14] years old) within each neighborhood (Fig 3A, Fig 3B and Fig 3C). There was no significant difference in specific IgG responses according to the age group whatever the neighborhood (*p*>0.05). However, highly significant variation in the specific IgG level was observed between the three study sites in children aged [1-5] and [6-10] years-old (*p*=0.001, Table 2). Indeed, as observed for the whole studied population, the specific IgG responses were higher in Bromakote compared to the two other neighborhoods in these youngest age groups. In contrast, no significant differences of specific IgG level were observed between the three neighborhoods in the [11-14] age group.

**Figure 3.**
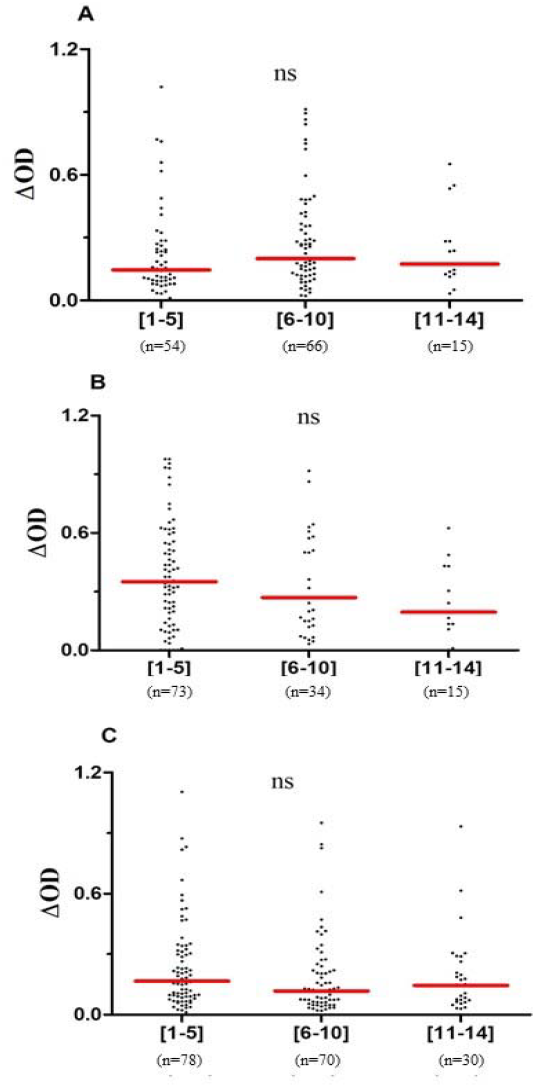
IgG responses to *Aedes aegypti* the Nterm-34 kDa salivary peptide according to age. Each dot represents the individual IgG response (ΔOD) against the *Ae. aegypti* Nterm-34 kDa salivary peptide and the horizontal bar indicates the median value of specific IgG levels for each age group. Results are presented according to the 3 studied sites A: Anoumabo, B: Bromakote and C: Petit-Bassam. The number of processed sera is indicated below (*n*).

### Entomological indices

Only three mosquito species were collected during the short rainy season across the three neighborhoods, with *Aedes* aegypti being the only species identified within the *Aedes* genus (Table 3). Three entomological methods were employed, each targeting different mosquito life stages. The dipping method was used for larval surveys, while ovitraps (followed by rearing) and sticky traps were used to assess adult mosquito populations.

**Table 3.**
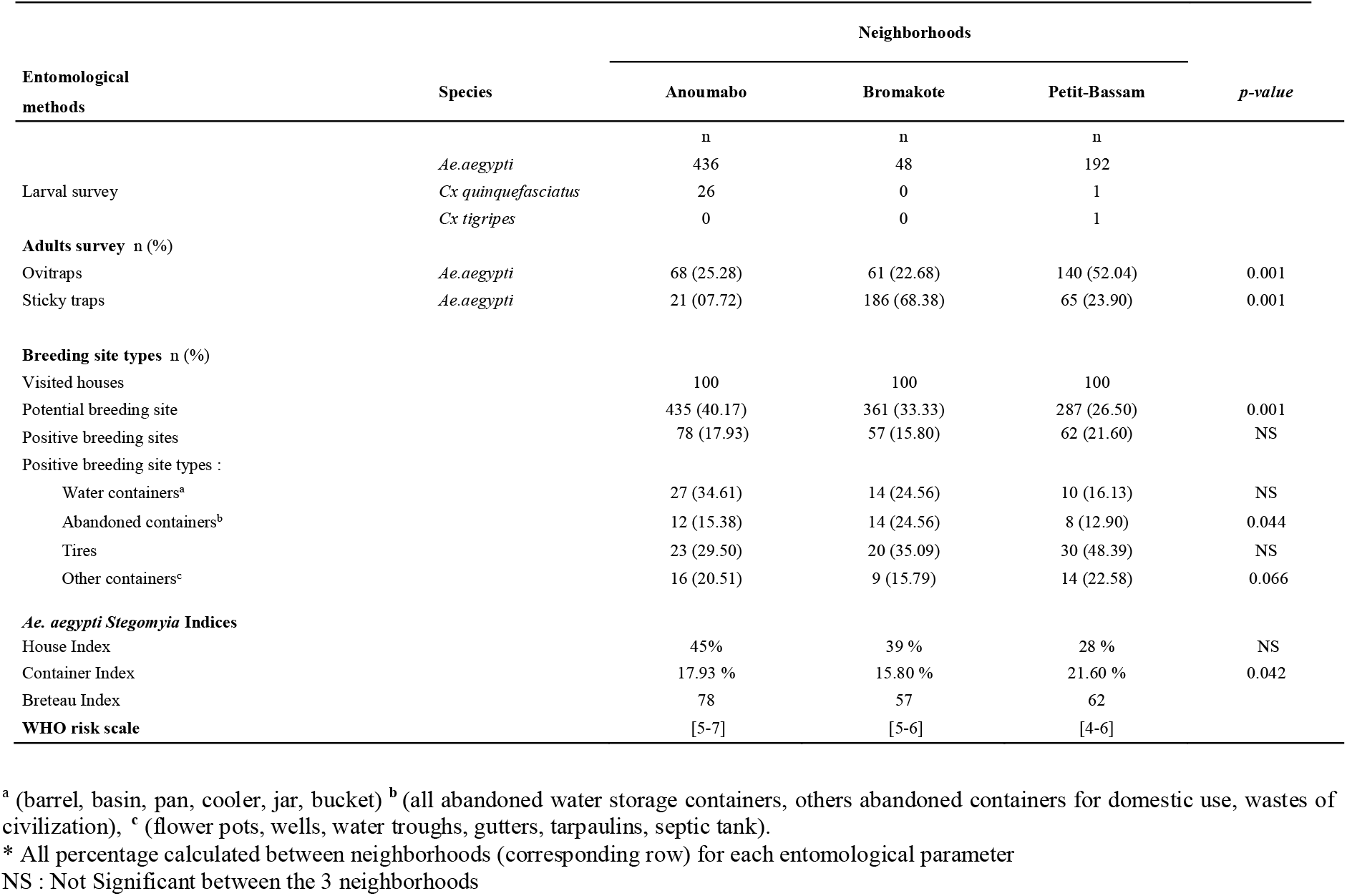
Entomological data in the study sites

The comparison of results using the ovitraps showed a significant difference in the *Ae. aegypti* proportion between the 3 neighborhoods (*p*=0.001), with the highest proportion observed in Petit-Bassam. In contrast, with sticky traps, a significantly higher proportion of adult mosquitoes was observed in Bromakote (68.38 % compared to the two other sites, *p*=0.001). In visited houses, 1083 potential breeding sites were counted (435 in Anoumabo, 361 in Bromakote, and 287 in Petit-Bassam). The proportions of potential breeding sites were significantly higher in Anoumabo compared to the two other neighborhoods (*p*=0.001). The type of positive breeding sites appeared also different according to the neighborhoods with a higher proportion of water storage containers in Anoumabo (NS), abandoned containers in Bromakote (*p*<0.05), and tires in Petit-Bassam (NS). Finally, the major *Stegomyia* indices were calculated. The HI and BI parameters appeared higher in Anoumabo but the difference was not significant between the three sites. In contrast, CI was different according to the studied sites and higher in Petit-Bassam (*p*<0.05). The WHO risk scale was thus calculated and 5-7, 5-6 and 4-6 scores were obtained in Anoumabo, Bromakote and Petit-Bassam, respectively. These scores suggested a similar risk between neighborhoods even if a slightly higher risk was noticed in Anoumabo as observed for BI.

## Discussion

In the present study, the quantitative and specific IgG responses to the *Aedes aegypti* Nterm-34 kDa salivary peptide were measured in order to evaluate and compare the level of exposure to *Aedes aegypti* bites in children from three neighborhoods of Abidjan city.

Firstly, the immunological data showed individual heterogeneity of IgG response to the *Aedes* salivary peptide within each studied neighborhood. This suggests that children were differently exposed to *Ae. aegypti* bites in each studied neighborhood. This heterogeneous exposure *to Aedes* appears to be closely associated with the urban context, where breeding sites are likely to be highly diverse (Elanga Ndille *et al*., 2012; Sagna *et al*., 2019). In addition, various individual factors (such as personal protection measures or attractiveness to mosquitoes) and household characteristics could contribute to these results.

Secondly, the major result of this study is that IgG levels to *Aedes aegypti* Nterm-34 kDa salivary peptide varied among populations living in different neighborhoods within the same city. This suggests varying levels of human exposure to *Aedes* vectors across the neighborhoods of Abidjan city. The influence of some social practices or environmental factors (i.e., the mode of water storage containers) may explain these differences in *Aedes* exposure, as previously reported (Sagna *et al*., 2018). In addition, some entomological parameters assessed in the present study may be linked to the observed variations between neighborhoods, including adult *Aedes* caught by the sticky traps, the types of breeding sites, and particularly abandoned water storage containers. As observed with the “adult” entomological methods (capture of adult), the specific IgG level was significantly higher in Bromakote compared to other neighborhoods. The immunological results suggest that children in Bromakote were more exposed to *Ae. aegypti* bites than those living in Anoumabo and Petit-Bassam. Interestingly, specific IgG levels appeared to be associated with only some entomological indicators. This discrepancy may be attributed to the entomological methods used in this study, as previously reported (Sagna *et al*., 2018). One hypothesis is that some entomological parameters may not be well-suited to the African urban context (WHO, 2016; Nascimento *et al*., 2020).

Thirdly, some environmental factors (waste management, water storage) and social practices (household type and construction materials) were compared between the studied neighborhoods. The objective was to identify the factors that could explain the site-dependent levels of *Aedes* exposure. Indeed, *Ae. aegypti* mosquitoes are known to exploit any abandoned containers that can hold water, creating ideal conditions for egg laying (Chareonviriyaphap *et al*., 2003; Hiscox *et al*., 2013). Unfortunately, no data on a potential association between socio-environmental level and specific IgG response was available at the individual level. These gaps could not allow statistical analyzes to be carried out. Our study represented an initial approach at the population and descriptive levels.

The active and permanent circulation of the dengue virus in West Africa is maintained by *Aedes* vectors, which find adequate breeding conditions for their development, as reported in Ouagadougou, Burkina Faso (Fournet *et al*., 2016). The present study highlighted that water containers, abandoned containers or tires could be the major breeding sites according to the studied neighborhoods. In addition, the proportion of children living in an apartment was higher in Bromakote compared to other studied neighborhoods. Although the number of apartments was low in the present study, one hypothesis is that this type of housing may also contribute to increased *Aedes* exposure, as previously suggested (Poinsignon *et al*., 2019). Indeed, the presence of potential breeding sites in apartments and/or the higher concentration of human population could play a significant role in this type of housing, compared to individual households. Nevertheless, further studies are needed to support this hypothesis, and particularly at the individual and/or household levels.

The IgG level to *Aedes aegypti* Nterm-34 kDa salivary peptide was also analyzed according to 3 age groups ([1-5], [6-10], and [11-14] years old) and the results showed heterogeneity in specific IgG levels within each age group. Children appeared therefore to be differently exposed to *Ae. aegypti* bites in each group. The youngest children were differently exposed to *Aedes* between the studied neighborhoods. At this young age (1 to 5 years old), children typically stay at home, and could be therefore exposed to the *Aedes* mosquito which breeds in the abandoned containers in the compound and have daily biting activities (Sagna *et al*., 2019). Those attending school and aged between 6 to 14 years could be also bitten by *Ae. aegypti* in the schoolyard or the classroom, as previously observed (Garcia-Rejon *et al*., 2011). Nevertheless, specific IgG responses remained higher in Bromakote compared to the other sites, but only in children of [1-5] and [6-10] year’s age groups, as observed in the entire studied population. It suggests that the younger children (<10 years old) could represent an appropriate target population for identifying the heterogeneity of human exposure to *Aedes* bites in urban environments. Altogether, the results indicated that children of all ages were at risk of arboviral virus transmission in urban settings, which should be considered for implementation of control strategies.

Overall, the present study highlights that the IgG level to *Ae. aegypti* Nterm-34 kDa salivary peptide could represent a relevant tool in the African urban context. It can be used to identify the neighborhoods at risk of exposure to *Ae. aegypti* bites and, consequently, at risk of arbovirus transmission (Elanga Ndille *et al*., 2014; Sagna *et al*., 2018 and Sagna *et al*., 2019). Nevertheless, the present study presents some limitations. Indeed, socio-environmental data were not directly collected from the household of sampled children which would have been relevant for integrating these variables into a statistical model to assess their actual impact on specific IgG level.

In conclusion, this study highlights that IgG level against *Ae. aegypti* Nterm-34 kDa salivary peptide represents a reliable indicator for evaluating the heterogeneity of children exposure to *Ae. aegypti* bites both at individual and population levels in African urban context. It could also provide an alternative approach to establish new criteria for measuring the efficacy of specific vector control strategies in urban settings, enabling more relevant monitoring of the programmes developed by public health authorities in African regions.

## Data Availability

All data produced in the present study are available upon reasonable request to the authors

## Appendices

### Ethics statement

The present study followed the ethical principles recommended by the Edinburgh revision of the Declaration of Helsinki. The protocol was approved by the Ethics Committee of Côte d’Ivoire, Ministry of Health (June 2014; N° 41/MSLS/CNER-dkn). Written informed consent of all the parents or legal guardians of children who participated in this study was obtained before inclusion.

## Acknowledgements

We gratefully acknowledge the populations of Anoumabo, Bromakote, and Petit-Bassam, especially householders and guardians of children, for their kind support and collaboration.

## Funding

This research was integrated into the ARBORISK multidisciplinary study funded through the C2D program (Contrat de Désendettement et de Développement; 2013). BZH Zamble was supported by a PhD fellowship provided by the Méditerannée Infection Foundation. DD Zoh was supported by a fellowship from the IRD.

## Conflict of interest disclosure

The authors declare that they comply with the PCI rule of having no financial conflicts of interest in relation to the content of the article.

## Data, script, code, and supplementary information availability

All data are available as supplementary material at https://www.medrxiv.org/content/10.1101/2025.03.25.25324614v3.supplementary-material.

### Legend of supplementary data

**ID** (column A) : anonymous code of individual

**Ag+** : individual OD value in one well with salivary antigen

**mean Ag+** : mean of the individual OD value in both wells with salivary antigen

**Ag -** : individual OD value in a blank well containing no antigen

**ΔOD** = OD value of mean Ag+ - OD value of Ag-

**SD**: Standard Deviation of duplicate of OD value of Ag+ wells

**CV**: Coefficient of Variation between duplicate value of Ag+ wells

## Notes

### Competing Interest Statement

The authors have declared no competing interest.

### Funding Statement

This research was integrated into the ARBORISK multidisciplinary study funded through the C2D program (Contrat de Desendettement et de Developpement; 2013). BZH Zamble was supported by a PhD fellowship provided by the Mediterannee Infection Foundation. DD Zoh was supported by a fellowship from the IRD.

### Author Declarations

The protocol was approved by the Ethics Committee of Cote d'Ivoire, Ministry of Health (June 2014; Numero 41/MSLS/CNER-dkn).

### Summary of Updates

All data are available as supplementary material and text of legends of supplementary data have been added in corresponding section of manuscript

